# Protocol and statistical analysis plan for the Antibiotic Choice On ReNal outcomes (ACORN) randomized clinical trial

**DOI:** 10.1101/2022.10.06.22280789

**Authors:** Edward T. Qian, Jonathan D. Casey, Adam Wright, Li Wang, Justin K. Siemann, Mary Lynn Dear, Joanna L. Stollings, Brad Lloyd, Kevin P. Seitz, George Nelson, Patty Wright, Edward D. Siew, Brad Dennis, Jesse O. Wrenn, Jonathan W. Andereck, Wesley H. Self, Matthew W. Semler, Todd W. Rice, the Vanderbilt Learning Healthcare System Platform Investigators

**Affiliations:** Vanderbilt University Medical Center, Department of Medicine, Division of Allergy, Pulmonary and Critical Care Medicine; Vanderbilt University Medical Center, Department of Bioinformatics; Vanderbilt University School of Medicine, Department of Biostatistics; Vanderbilt University Medical Center, Vanderbilt Institute for Clinical and Translational Research; Vanderbilt University Medical Center, Department of Pharmaceutical Services; Vanderbilt University Medical Center, Department of Emergency Medicine.; Vanderbilt University Medical Center, Department of Medicine, Division of Infectious Disease; Vanderbilt University Medical Center, Department of Medicine, Division of Nephrology and Hypertension; Vanderbilt University Medical Center, Department of Surgery

**Keywords:** Antibacterial Agents, Sepsis, Acute Kidney Injury, Delirium

## Abstract

**Introduction:** Antibiotics are time-critical in the management of sepsis. When infectious organisms are unknown, patients are treated with empiric antibiotics to include coverage for gram-negative organisms, such as anti-pseudomonal cephalosporins and penicillins. However, in observational studies some anti-pseudomonal cephalosporins (e.g. cefepime) is associated with neurologic dysfunction while the most common anti-pseudomonal penicillin (piperacillin-tazobactam) is associated with acute kidney injury. No randomized control trials have compared these regimens. This manuscript describes the protocol and analysis plan for a trial designed to compare the effects of anti-pseudomonal cephalosporins and anti-pseudomonal penicillins among acutely ill patients receiving empiric antibiotics.

**Methods and Analysis:** The <u>A</u>ntibiotic <u>C</u>hoice <u>O</u>n <u>R</u>e<u>N</u>al outcomes (ACORN) trial is a prospective, single-center, non-blinded randomized trial being conducted at Vanderbilt University Medical Center. The trial will enroll 2,500 acutely ill adults receiving gram-negative coverage for treatment of infection. Eligible patients are randomized 1:1 to receive cefepime or piperacillin-tazobactam upon first order entry of a broad-spectrum antibiotic covering gram-negative organisms. The primary outcome is the highest stage of acute kidney injury and death occurring between enrollment and 14 days after enrollment. This will be compared between patients randomized to cefepime and randomized to piperacillin-tazobactam using an unadjusted proportional odds regression model. The secondary outcomes are Major Adverse Kidney Events through day 14 and number of days alive and free of delirium and coma in 14 days after enrollment. Enrollment began on November 10, 2021 and is expected to be completed in December 2022.

**Ethics and Dissemination:** The trial was approved by the Vanderbilt University Medical Center institutional review board with a waiver of informed consent. Results will be submitted in a peer-reviewed journal and presented at scientific conferences.

**Trial Registration:** This trial was registered with ClinicalTrials.gov (NCT05094154) on October 26, 2021, prior to enrollment of the first patient on November 10, 2021.

**Strengths and Limitations:**

- This ongoing pragmatic trial will compare the effects of cefepime vs piperacillin-tazobactam on acute kidney injury and death among acutely ill adults receiving gram-negative antibiotic therapy in the Emergency Department or Intensive Care Unit.
- Strengths: Broad eligibility criteria, inclusion of a range of indications for antibiotic therapy, and use of the electronic health record to screen for eligible patients and facilitate delivery of the assigned intervention will increase the external validity of the findings
- Limitations: After concealed randomization, patients, clinicians, and investigators are unblinded to study group assignment. Because urine output is not systematically available across all care, the outcome of AKI is based on creatinine measurements.

## Introduction

Antibiotics are necessary for management of patients with sepsis^1^ but can cause unintended adverse effects on organ function^2^. Since specific organisms causing infection is often unknown, empiric broad-spectrum antibiotics are commonly prescribed. For patients at risk for resistant organisms, common regimens include gram-positive coverage (i.e. vancomycin) and gram-negative coverage with an anti-pseudomonal cephalosporin or penicillin, predominantly cefepime or piperacillin-tazobactam^1^.

Because the medications are considered to have comparable anti-pseudomonal activity, discussion surrounding choice has focused on adverse effects. Some observational studies have reported an association between receipt of piperacillin-tazobactam and acute kidney injury (AKI)^3,4^, particularly among patients receiving vancomycin^5–7^. Other studies have shown no relationship between piperacillin-tazobactam and AKI^8,9^. AKI is common during hospitalization^10^, and there are many potential contributors including isotonic fluids^11,12^, medications, and acute illnesses like sepsis^13,14^.

Similarly, an association between cephalosporins and neurotoxicity manifesting as delirium and coma has been observed.^15–17^ Delirium, acute brain dysfunction characterized by fluctuations in mental status, inattention, altered consciousness, and disorganized thinking,^18^ is also a common complication of hospitalization. In Intensive Care Unit (ICU) populations, delirium is predictive of mortality, prolonged length of stay, and long-term cognitive impairment^19,20^. The incidence of cephalosporin-induced neurotoxicity is unknown but has been reported to increase in-hospital mortality^21^.

A randomized controlled trial would overcome limitations of observational data, but none are known to exist^22^. Rigorous, high-quality evidence assessing risk of AKI and neurotoxicity after exposure to anti-pseudomonal antibiotics would have potential to change care received by thousands of acutely ill adults annually. To address the lack of available evidence, we are conducting a prospective, randomized trial comparing anti-pseudomonal cephalosporins and anti-pseudomonal penicillins among acutely ill adults in the Emergency Department (ED) or ICU.

## Methods and Analysis

This manuscript was written in accordance with Standard Protocol Items: Recommendations for Interventional Trials (SPIRIT) guidelines (Table 1, Supplement 1)^23^. The Learning Healthcare System (LHS) Platform at Vanderbilt University Medical Center conducts research studies using a unique model that leverages pragmatic, randomized, controlled clinical trials embedded within usual care^24^. The LHS Platform is composed of stakeholders from across the enterprise and supports projects in both the pediatric and adult inpatient and outpatient settings (Supplement 2). LHS Platform studies focus on comparative effectiveness, implementation science, and programmatic evaluation approaches^25,26^.

**Table 1.** Standard Protocol Items: Recommendations for Interventional Trials (SPIRIT) checklist. Enrollment, interventions, and assessments. CEF, Cefepime; PTZ, piperacillin-tazobactam; EHR, Electronic health record;

|  | STUDY PERIOD |  |  |  |  |
| --- | --- | --- | --- | --- | --- |
|  | Eligibility Screen | Randomization & Allocation | Post-allocation |  | Final Outcome Assessment |
| TIMEPOINT | Order entry for CEF or PTZ | EHR enrollment advisor | 7 days after enrollment | 14 days after enrollment | Discharge or 28 days after enrollment |
| **ENROLLMENT:** |  | X |  |  |  |
| EHR-based inclusion criteria screening | X |  |  |  |  |
| Manual screening for exclusion criteria by treating clinicians | X |  |  |  |  |
| Allocation |  | X |  |  |  |
| **INTERVENTIONS:** |  |  |  |  |  |
| *Cefepime* |  | X | X |  |  |
| *Piperacillin-tazobactam* |  | X | X |  |  |
| **ASSESSMENTS:** |  |  |  |  |  |
| *Baseline variables* | X | X |  |  |  |
| *Adverse events* |  | X | X | X | X |
| *Primary and secondary outcome* |  | X | X | X |  |
| *Exploratory outcomes* |  |  | X | X | X |

### Study Design

The <u>A</u>ntibiotic <u>C</u>hoice <u>O</u>n <u>R</u>e<u>N</u>al outcomes (ACORN) trial is a pragmatic, single-center, unblinded, parallel-group, randomized trial comparing anti-pseudomonal cephalosporins to anti-pseudomonal penicillins among acutely ill adults receiving gram-negative antibiotics in the ED and ICU. At this center, the predominant anti-pseudomonal cephalosporin is cefepime and the predominant anti-pseudomonal penicillin is piperacillin-tazobactam. The primary outcome is highest stage of AKI or death in 14 days. The trial protocol was approved by the institutional review board at Vanderbilt University Medical Center and registered prior to initiation of enrollment (NCT05094154). An independent data and safety monitoring board (DSMB) monitors the progress and safety of the trial.

### Study Population

#### Inclusion criteria

1. Age ≥ 18 years old
2. Located in a participating ED or ICU
3. Less than 12 hours between presentation to study hospital
4. Treating clinician initiating an order for an anti-pseudomonal cephalosporin or anti-pseudomonal penicillin

#### Exclusion criteria

1. Known receipt of > 1 dose of an anti-pseudomonal cephalosporin or anti-pseudomonal penicillin during the last 7 days
2. Current documented allergy to cephalosporins or penicillin
3. Known to be a prisoner
4. Treating clinicians feel that either an anti-pseudomonal cephalosporin or anti-pseudomonal penicillin is required or contraindicated for the optimal treatment of the patient, including for more directed antibiotic therapy against known prior resistant infections or suspected sepsis with an associated central nervous system infection

### Screening and Enrollment

When a treating clinician in a participating ED or ICU initiates an order for cefepime or piperacillin-tazobactam for a patient who meets all inclusion criteria, a clinical decision support (CDS) tool will 1) inform the provider of the study, 2) query the provider regarding the presence of any exclusion criteria, and if none are present, 3) enroll and randomize the patient. For patients who meet an exclusion criterion, the reason is recorded (Fig. 1).

**Figure 1.**
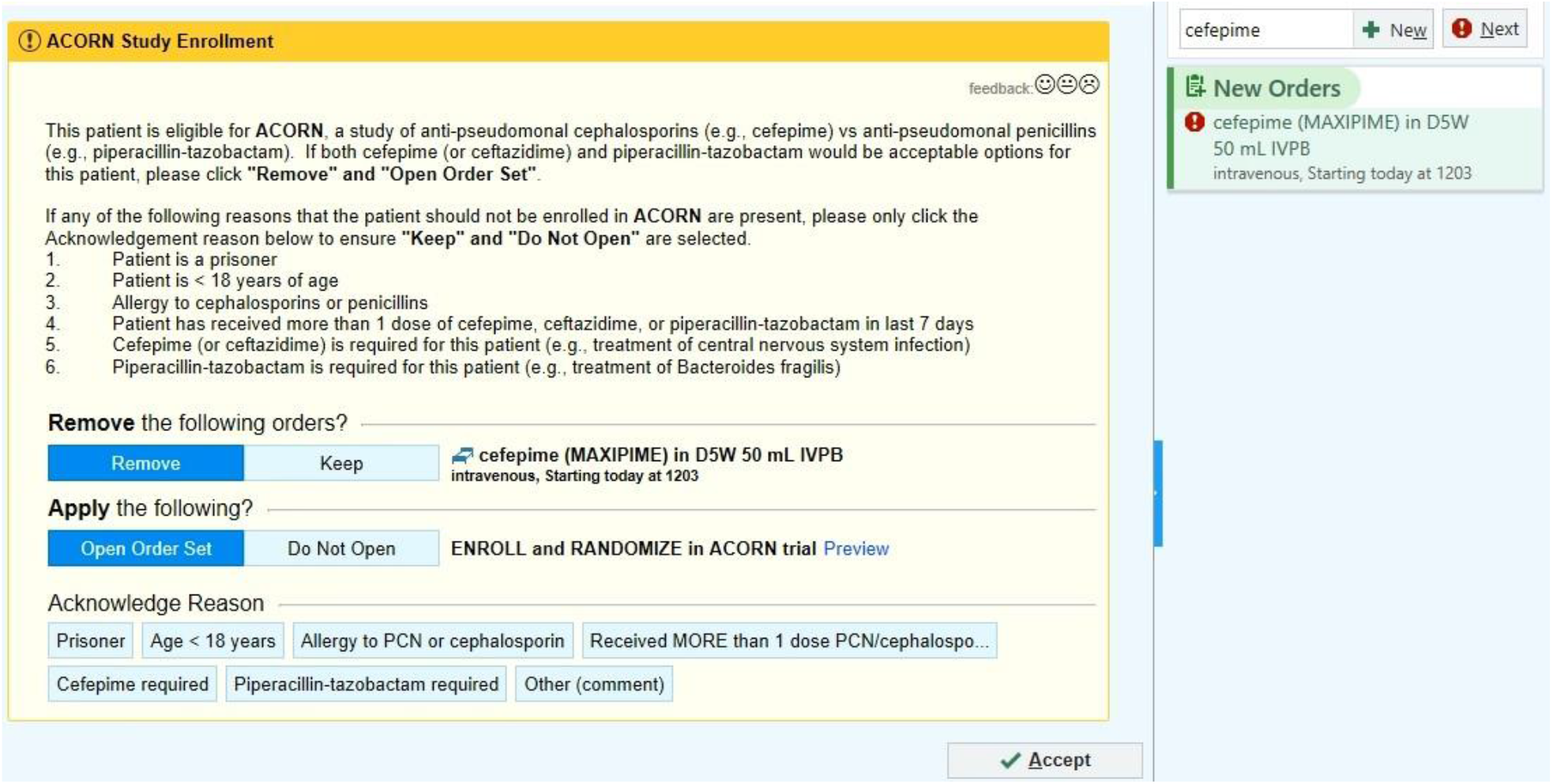
Electronic health record-based enrollment advisor

### Analysis population

The primary goal is the exploration of uncommon safety effects, therefore the population for primary analysis will include all patients who were randomized and received at least one dose of either study drug in the 168 hours (7 days) after randomization. A sensitivity analysis will include all patients randomized, including those who never received either cefepime or piperacillin/tazobactam.

### Randomization and Treatment Allocation

Study group assignments are generated by computerized randomization in a 1:1 ratio of intervention to control. Study group assignment will remain concealed until the team has confirmed the patient does not meet any exclusion criteria and the patient has been enrolled. The CDS tool will advise the clinician of group assignment following randomization.

### Study Interventions

For patients assigned to cefepime, the CDS tool will guide providers to intravenous cefepime. For patients assigned to piperacillin-tazobactam, the CDS tool will guide providers to intravenous piperacillin-tazobactam. The CDS tool will display a standardized table of dose by glomerular filtration rate, but decisions regarding dose, frequency, and duration will be the treating clinician’s discretion.

In the 168 hours (7 days) following enrollment, any new order for cefepime or piperacillin-tazobactam will open a CDS tool, which displays group assignment and allows clinicians to (re)order the assigned antibiotic or provide a reason for ordering the non-assigned antibiotic. Similarly, if the assigned antibiotic is discontinued, the CDS tool will solicit a rationale for discontinuation, including antibiotic tailoring, newly apparent allergy to either cephalosporins or penicillins, or clinician preference.

The CDS tool influences only the choice of the initial anti-pseudomonal antibiotic. Treating clinicians determine concurrent administration of other antibiotics (e.g., vancomycin, metronidazole), duration of therapy, escalation, de-escalation, approach to source control, and use of culture and laboratory data to modify antibiotic therapy.

### Data Collection

Trial personnel will monitor for adverse events daily and will record the following data elements at the time of enrollment by manual review of the health record (Supplement 3 and 4):

- Presence of sepsis, defined by Sepsis-3 criteria^27^
- Transplant recipient status
- Receipt of renal replacement therapy (RRT) prior to enrollment
- Presumed source of infection organized into groups based on previously published data^28^

All other data will be obtained using electronic exports from the health record. The following variables are collected:

1. Collected at baseline: Age, sex, height, weight, body mass index, race, Sequential Organ Failure Assessment (SOFA) score, active medical problems at the time of admission, active comorbidities, comorbidities and medications known to increase risk of kidney or neurologic injury at enrollment, mean arterial pressure and vasopressor use prior to antibiotic receipt, analgesia and sedation use prior to antibiotic receipt, admission to ICU vs ward
2. Collected from randomization to hospital discharge: Mean arterial pressure and vasopressor use, pH, PaO2, PaCO2, respiratory rate, heart rate, oxygen saturation, temperature, lactic acid, elements of a basic metabolic panel, magnesium, elements of a complete blood count, antibiotic receipt, all microbiologic culture data, Confusion Assessment Method for the ICU (CAM-ICU), Richmond Agitation Sedation Score (RASS), Glasgow Coma Scale (GCS), mechanical ventilation status and variables related to ventilation, nephrology consultation, receipt of new RRT, indications for new RRT among patients who received new RRT, medications known to increase risk of kidney injury, medications known to increase risk of neurologic injury, admitting team, date of admission, days spent in the ICU, days spent in the hospital, date of intubation(s) and extubation(s), date of discharge, and date of death.

### Primary Outcome

The primary outcome will be a combination of the highest stage of AKI and death between randomization and day 14. The stages of AKI are defined using creatinine measurements and the “Kidney Disease: Improving Global Outcomes (KDIGO)”^29^ criteria. The score will range from 0 (best value) to 4 (worst value):

0 = No AKI

1 = Stage 1 AKI (Creatinine increased by 1.5-1.9 times baseline OR increase by >= 0.3 mg/dL)

2 = Stage 2 AKI (Creatinine increased by 2.0-2.9 times baseline)

3 = Stage 3 AKI (Creatinine increased by >= 3.0 times baseline OR increase to >= 4.0 mg/dL OR New RRT)

4 = Death

“Baseline” creatinine values are defined as the lowest prior creatinine values from three different timepoints: the pre-illness creatinine value, the peri-enrollment creatinine value, and the lowest prior on-study creatinine value as defined below. Death is defined as mortality from any cause occurring prior to or on the end of study day 14, censored at hospital discharge. RRT is defined as receipt of RRT at any point between randomization and the end of study day 14, censored at hospital discharge. Patients who are receiving RRT prior to enrollment can only experience values of 0 (patient did not die) or 4 (patient died) because they are ineligible to experience changes in creatinine or new receipt of RRT that define levels 1 through 3.

Patients’ pre-illness creatinine will be defined as the lowest serum creatinine between 12 months and 24 hours prior to enrollment. For patients for whom a value is unavailable, a pre-illness creatinine value will be estimated using a previously-described three-variable formula [creatinine = 0.74 – 0.2 (if female) + 0.08 (if African American) + 0.003 × age (in years)]^30^. There are no validated estimations of creatinine without race but we will evaluate the effect of social constructs by fitting models both with and without race in a sensitivity analysis.

Patients’ peri-enrollment creatinine will be defined hierarchically using the creatinine value closest to enrollment in: (first) the 24 hours prior to enrollment, if available, and (second) the six hours after enrollment (if no value is available prior to enrollment). Prevalent AKI, or AKI that is present on admission and unrelated to the study intervention, will be defined by comparing the peri-enrollment creatinine to the pre-illness creatinine.

On-study creatinine will be defined as any creatinine value occurring after both the time of enrollment and the time of the peri-enrollment creatinine value. On-study creatinine values will be used to identify incident AKI and calculate the stage of AKI for the primary outcome.

The primary outcome will be calculated as follows:

- Patients who survive without new RRT and do not experience an on-study creatinine value that is at least 0.3 mg/dL higher than the peri-enrollment value or any preceding on-study value, and whose on-study creatinine is never more than 1.5 times the peri-enrollment value or any preceding on-study value, will be considered not to have experienced incident AKI and will receive a value of 0 for the primary outcome.
- Among patients who survive and experience AKI (have on-study creatinine value that is at 1.5 times or at least 0.3 mg/dL higher than the peri-enrollment value or any preceding on-study value), the score for the primary outcome will be determined by the stage of AKI and classified as follows:
  a. A value of 1 if the highest on-study creatinine after qualifying for AKI is less than 2.0 times the lowest of the pre-illness creatinine value, the peri-enrollment creatinine value, and the lowest prior on-study creatinine value (baseline creatinine);
  b. A value of 2 if the highest on-study creatinine after qualifying for AKI is at least 2.0 times and less than 3.0 times the lowest of the pre-illness creatinine value, the peri-enrollment creatinine value, and the lowest prior on-study creatinine value (baseline creatinine); and
  c. A value of 3 if the highest on-study creatinine after qualifying for AKI is at least 3.0 times the lowest of the pre-illness creatinine value, the peri-enrollment creatinine value, and the lowest prior on-study creatinine value (baseline creatinine), with a maximum creatinine above 4 mg/dL, or receive new RRT
- Patients who die will receive a value of 4

The mechanisms and extent of AKI with antibiotic exposure are not well understood. The primary outcome window of 14 days was chosen as it was felt to capture the period most likely to be affected by controlling antibiotics choice for 168 hours (7 days).

### Secondary Outcomes

We have prespecified two secondary outcomes. Major Adverse Kidney Events within 14 days (MAKE14), is the composite outcome of death within 14 days, new RRT within 14 days, or stage 2 or higher AKI at day 14, according to KDIGO creatinine criteria. The second is number of days alive and free of delirium and coma in the 14 days after enrollment (Delirium and Coma-Free Days to day 14). Delirium is defined as a positive assessment on the CAM-ICU^31^ and coma is defined as a RASS of −4 or −5^32^ at any point during that study day.

### Exploratory Outcomes

- Exploratory Renal Outcomes
  - Major Adverse Kidney Events within 28 days (MAKE28)
  - Highest stage of AKI or death between randomization and day 7
  - Stage 2 or higher AKI as defined in the KDIGO criteria for creatinine level within 14 days and 28 days after enrollment
  - New receipt of RRT within 14 days and 28 days after enrollment
  - Days alive and free of RRT during 14 days and 28 days after enrollment
  - Highest creatinine level within 28 days after enrollment
  - Change from pre-illness creatinine to the highest creatinine level within 28 days after enrollment
  - Final creatinine level before hospital discharge at 28 days
  - Ongoing receipt of RRT at hospital discharge or 28 days
  - Nephrology consultation
- Exploratory Neurologic Outcomes
  - Worst GCS score during the 7 days, 14 days, and 28 days after enrollment
  - Delirium and Coma-Free Days in the 28 days after enrollment
- Exploratory Clinical Outcomes
  - ICU-free, Hospital-free, Ventilator-free, and Vasopressor-free days in the 28 days after enrollment
  - 14-day mortality
  - 28-day mortality
  - Disposition of patients admitted from the ED (ward vs ICU)
  - Escalation of antibiotics defined by subsequent receipt of meropenem, meropenem-vaborbactam, imipenem, imipenem-relebactam, cefiderocol, ceftazidime-avibactam, ceftolozane-tazobactam, tigecycline, amikacin, tobramycin, gentamicin.

Definition of Supportive Therapy-Free days is available in Supplement section 5.

### DSMB and Interim Analysis

A DSMB composed of experts in critical care medicine, infectious disease, and biostatistics has overseen the design of the trial and is monitoring its conduct (Supplement 6). The DSMB conducted a single interim analysis, prepared by the study biostatistician, at the anticipated halfway point of the trial after enrollment of 1025 patients. The meeting was held on July 14, 2022, and the DSMB recommended continuing the trial to completion without alteration. The stopping boundary for efficacy was prespecified as p-value for the difference between groups in the primary outcome of 0.001 or less. Given current use of both interventions as a part of usual care, there was no stopping boundary for futility. Use of a conservative Haybittle-Peto boundary for efficacy will allow the final analysis to be performed using an unchanged level of significance (P = 0.05). The DSMB retains the authority to recommend stopping the trial at any point, request additional data or interim analyses, or request modifications of study protocol to protect patient safety.

### Sample Size Estimation

As specified in the initial trial protocol, at the time of the single, planned interim analysis the DSMB oversaw a re-estimation of the planned sample size. Because [1] concurrent receipt of vancomycin has been hypothesized to be in the proposed mechanistic pathway between receipt of an anti-pseudomonal penicillin and AKI and [2] approximately 75% of patients in the trial concurrently receive vancomycin, the sample size was increased by 25% from 2,050 to 2,500 patients. The increase in sample size ensures the number of patients receiving concurrent vancomycin will be approximately 2,050, consistent with the original sample size estimation. Assuming a two-sided alpha of 0.05 and a distribution of the primary outcome with approximately 70% of patients experiencing no AKI, 10% of patients experiencing stage I AKI, 7% of patients experiencing stage II AKI, 7% of patients experiencing stage III AKI, and 6% of patients experiencing death, we calculated that enrollment of a total of 2,500 patients would provide 92% statistical power to detect an odds ratio of 0.75 in the primary analysis.

### Statistical Analysis Principles

Analyses will be conducted following reproducible research principles using R (R Foundation for Statistical Computing, Vienna, Austria)^33^. Continuous variables will be reported as mean ± SD or median and IQR; categorical variables will be reported as frequencies and proportions. As a randomized controlled trial, there will be no comparison of baseline characteristics. A two-sided p-value of < 0.05 will be used to indicate statistical significance; with just one primary outcome, no adjustment for multiplicity will be made. Two secondary outcomes are specified, one safety outcome for each treatment regimen. Since hypothesized safety concerns are independent, we will not adjust our secondary outcomes for multiplicity. For all other outcomes, emphasis will be placed on magnitude of differences between groups rather than statistical significance.

### Main Analysis of the Primary Outcome

The main analysis will be an unadjusted, intention-to-treat comparison of the primary outcome between patients randomized to receive anti-pseudomonal cephalosporins versus anti-pseudomonal penicillins who received at least one dose of a study drug using a proportional odds regression model. The unadjusted common odds ratio (cOR) with confidence intervals will be the primary treatment effect. If departures from the proportionality assumption are observed, then a partially proportional odds model will be constructed.

### Secondary Analyses of the Primary Outcome

#### Multivariable modeling to account for covariates

To account for participants’ baseline status, we will include covariates in the proportional odds regression model. The following prespecified baseline covariates will be considered: age; sex; peri-enrollment creatinine; receipt of RRT prior to enrollment; receipt of vasopressors; receipt of mechanical ventilation; SOFA score; presumed source of infection; enrollment location (ED vs ICU). Source of infection will be categorized as: lung, intra-abdominal (perforated viscus, ischemic bowel, cholecystitis/cholangitis, peritonitis/abscess/small bowel obstruction, *Clostridium difficile* colitis, spontaneous bacterial peritonitis, pancreatitis, enterocolitis/diverticulitis, other intra-abdominal infections), urinary (pyelonephritis, obstructive urinary tract infection), skin and soft tissue (cellulitis/abscess/necrotizing fasciitis/decubitus ulcer, surgical site infection), other (bone/joint, primary blood stream infection, intravascular catheter, disseminated infection, central nervous system infection, endocarditis, other), and unknown.

#### Effect Modification

We will test the interaction between the treatment effect of anti-pseudomonal cephalosporins vs anti-pseudomonal penicillins and baseline variables expected to modify effects of treatments on the outcomes. The effect modifiers will be tested one by one by including both the main effect and the interaction term in the adjusted model. Because this study is not formally designed or powered to test for interaction, a less conservative two-sided p value for the interaction term will be used, with values less than 0.10 considered suggestive of potential interaction and values less than 0.05 considered conclusive evidence. The following variables will be considered:

1. Location at randomization (ED vs ICU)
2. Presence of sepsis (meeting Sepsis-3 criteria) at randomization
3. Receipt of vancomycin (defined as an order for vancomycin in the 12 hours before or 6 hours after randomization)
4. Source of infection
5. AKI at randomization
6. CKD at randomization
7. Neutropenia at randomization
8. Admitting team (medicine vs surgical)

#### Sensitivity Analyses of the Primary Outcome

To assess robustness of findings, the main analysis of the primary outcome will be repeated in several alternative populations. First, we will include all patients who were identified by the CDS tool as meeting eligibility criteria, regardless of receipt of any doses of anti-pseudomonal cephalosporins or anti-pseudomonal penicillins. Second, many patients are initiated on antibiotics in the acute setting, which are stopped as a more likely cause for their illness becomes known (e.g. pulmonary embolism). We will repeat the main analysis restricted to the subset of patients that received more than 2 days (48 hours) of anti-pseudomonal therapy. Third, because race is a social construct, we will repeat the main analysis with those whose pre-illness creatinine was estimated by an equation without race as a factor. Fourth, to avoid uncertainty around the calculations of pre-illness creatinine, we will repeat the main analysis excluding patients with calculated pre-illness creatinine values. Because group assignment might influence recovery from prevalent AKI in a way that could affect calculations of the primary outcome, we will recalculate the primary outcome as a repeated measure assessed daily from randomization to day 14 using only the pre-illness creatinine as the baseline creatinine.

### Analysis of the Secondary Outcomes

Analysis of secondary outcomes will follow a similar framework to the primary analysis, with a systematic assessment of unadjusted models, adjusted models using the same set of covariates. The secondary outcome of MAKE14 will be compared between groups using a logistic regression model. Delirium and coma-free days to day 14 will be compared between groups using a proportional odds regression model, and will include the additional covariate of coma on enrollment for the adjusted model. Delirium and coma free days to day 14 will be assessed for effect modification using the same approach as the primary outcome replacing receipt of vancomycin with baseline coma.

### Analyses of Exploratory Outcomes

Exploratory outcomes will proceed using unadjusted analyses only, with presentation of effect sizes and confidence intervals as well as p-values. Continuous outcomes will be compared with the Mann-Whitney U test and difference in medians reported. For categorical variables, groups will be compared with the chi-square test or Fischer’s exact test as appropriate and results will be expressed as a difference in proportions or odds ratios, each with 95% confidence intervals.

### Handling of Missing Data

In the case that a patient is enrolled, never receives RRT, and is discharged alive without having a creatinine value measured following enrollment, the patient will be assumed to have no AKI. When data are missing for secondary or exploratory outcomes, complete-case analysis, excluding cases where data for the analyzed outcome are missing, will be performed. There will be no imputation of missing data for these outcomes. In adjusted analyses, missing data for covariates will be imputed using multiple imputations.

### Trial status

The ACORN trial is currently enrolling and started enrollment on November 10, 2021.

## Ethics and Dissemination

### Waiver of Informed Consent

Acutely ill patients for whom the provider is ordering broad-spectrum antibiotics in the ED or ICU are at significant risk for morbidity and mortality from their underlying illness. Most patients receiving empiric gram-negative antibiotics in routine clinical care receive either anti-pseudomonal cephalosporins or anti-pseudomonal penicillins. Any benefits or risks of these two approaches are experienced by patients receiving gram negative antibiotics in clinical care, outside the context of research. As a requirement for enrollment in the ACORN trial, the patient’s treating clinician must have made the decision to order either anti-pseudomonal cephalosporins or anti-pseudomonal penicillins as part of routine clinical care and affirmed that either would be a safe and reasonable approach for the patient (otherwise the patient is excluded). Therefore, making the decision between the two approaches randomly through study group assignment rather than by a provider who thinks either approach is safe and reasonable for the patient was proposed to pose no more than minimal incremental risk.

Obtaining informed consent for participation in the study would be impracticable. Receipt of antibiotics in sepsis is time sensitive. Each hour delay in antibiotics in patients with sepsis is associated with an increase in mortality^34^. Attempting to obtain prospective written informed consent from patients presenting to the ED or ICU during the interval between the placement of an order for empiric antibiotics and their administration risks delaying antibiotic delivery. Moreover, acutely ill patients with sepsis are commonly delirious or unconscious, and a legally authorized representative is not consistently present at the time of initiation of antibiotics. Because the trial determines only the choice of the initial anti-pseudomonal antibiotic and defers decisions regarding subsequent doses of antibiotics (e.g., duration of therapy, escalation, de-escalation) to treating clinicians, enrollment, trial group assignment, and the primary study procedure (administration of the assigned antibiotic) commonly occurs within 1 hour of meeting eligibility criteria.

Because the study was expected to pose minimal risk and prospective informed consent was considered to be impracticable, a waiver of informed consent was requested and granted from the Vanderbilt University Medical Center IRB.

### Protocol Changes

ClinicalTrials.Gov will be updated with any amendments to the protocol as per SPIRIT guidelines (Supplement 7).

### Dissemination Plan

Trial results will be submitted to a peer-reviewed journal for consideration of publication and will be presented at one or more scientific conferences. Data will be made available following publication (Supplement 8).

## Conclusion

To allow for a clearer and more objective interpretation of trial results, this description delineates the ACORN trial methods and analysis prior to the conclusion of enrollment.

## Supporting information

Supplement for the ACORN SAP

## Data Availability

All data produce in the present work are contained in the manuscript

## Contributors

All study authors approved the final version of this manuscript. Study concept and design: E.T.Q., J.D.C., A.W., L.W., J.L.S., G.N., P.W., E.D.S., J.O.W., J.W.A., W.H.S., M.W.S., T.W.R.; Acquisition of data: E.T.Q., B.L., K.P.S.; Drafting of the manuscript: E.T.Q., J.D.C., M.W.S., T.W.R.; Critical revision of the manuscript for important intellectual content: E.T.Q., J.D.C., A.W., L.W., J.K.S., M.L.D, J.L.S., B.L., K.P.S., G.N., P.W., E.D.S., B.D., J.O.W., J.W.A., W.H.S., M.W.S., T.W.R.; Study supervision: M.W.S., T.W.R.

## Patient and Public Involvement

Patients or the public were not involved in the design, or conduct, or reporting, or dissemination plans of our research

