## Supplement for the ACORN SAP for "Protocol and statistical analysis plan for the Antibiotic Choice On ReNal outcomes (ACORN) randomized clinical trial"

#### Version 1

**Table of Contents**

1. SPIRIT 2013 Checklist
2. List of Vanderbilt Learning Healthcare System Platform Investigators
3. Data Auditing
4. Patient Privacy and Data Storage
5. Definition of Support free days
6. Data and Safety Monitoring Board Charter
7. Plan for communication of protocol changes
8. Public Access

#### 1. SPIRIT 2013 Checklist

SPIRIT

STANDARD PROTOCOL ITEMS: RECOMMENDATIONS FOR INTERVENTIONAL TRIALS

SPIRIT 2013 Checklist: Recommended items to address in a clinical trial protocol and related documents*

| Section/item | Item No | Description | Addressed on page number |
|---|---|---|---|
| **Administrative information** | | | |
| Title | 1 | Descriptive title identifying the study design, population, interventions, and, if applicable, trial acronym | 1 |
| Trial registration | 2a | Trial identifier and registry name. If not yet registered, name of intended registry | 4 |
| | 2b | All items from the World Health Organization Trial Registration Data Set | 1-4, 22 |
| Protocol version | 3 | Date and version identifier | 2 |
| Funding | 4 | Sources and types of financial, material, and other support | 22 |
| Roles and responsibilities | 5a | Names, affiliations, and roles of protocol contributors | 1, 22 |
| | 5b | Name and contact information for the trial sponsor | NA |
| | 5c | Role of study sponsor and funders, if any, in study design; collection, management, analysis, and interpretation of data; writing of the report; and the decision to submit the report for publication, including whether they will have ultimate authority over any of these activities | NA |

| | | | |
|---|---|---|---|
| | 5d | Composition, roles, and responsibilities of the coordinating centre, steering committee, endpoint adjudication committee, data management team, and other individuals or groups overseeing the trial, if applicable (see Item 21a for data monitoring committee) | __NA |

**Introduction**

| | | | |
|---|---|---|---|
| Background and rationale | 6a | Description of research question and justification for undertaking the trial, including summary of relevant studies (published and unpublished) examining benefits and harms for each intervention | __6 |
| | 6b | Explanation for choice of comparators | __6 |
| Objectives | 7 | Specific objectives or hypotheses | __6 |
| Trial design | 8 | Description of trial design including type of trial (eg, parallel group, crossover, factorial, single group), allocation ratio, and framework (eg, superiority, equivalence, noninferiority, exploratory) | __7 |

**Methods: Participants, interventions, and outcomes**

| | | | |
|---|---|---|---|
| Study setting | 9 | Description of study settings (eg, community clinic, academic hospital) and list of countries where data will be collected. Reference to where list of study sites can be obtained | __7 |
| Eligibility criteria | 10 | Inclusion and exclusion criteria for participants. If applicable, eligibility criteria for study centers and individuals who will perform the interventions (eg, surgeons, psychotherapists) | __9 |
| Intervention s | 11a | Interventions for each group with sufficient detail to allow replication, including how and when they will be administered | __10 |
| | 11b | Criteria for discontinuing or modifying allocated interventions for a given trial participant (eg, drug dose change in response to harms, participant request, or improving/worsening disease) | __10 |
| | 11c | Strategies to improve adherence to intervention protocols, and any procedures for monitoring adherence (eg, drug tablet return, laboratory tests) | _9-10 |

| | | | |
|---|---|---|---|
| | 11d | Relevant concomitant care and interventions that are permitted or prohibited during the trial | NA |
| Outcomes | 12 | Primary, secondary, and other outcomes, including the specific measurement variable (eg, systolic blood pressure), analysis metric (eg, change from baseline, final value, time to event), method of aggregation (eg, median, proportion), and time point for each outcome. Explanation of the clinical relevance of chosen efficacy and harm outcomes is strongly recommended | 11-15 |
| Participant timeline | 13 | Time schedule of enrolment, interventions (including any run-ins and washouts), assessments, and visits for participants. A schematic diagram is highly recommended (see Figure) | Table 1 |
| Sample size | 14 | Estimated number of participants needed to achieve study objectives and how it was determined, including clinical and statistical assumptions supporting any sample size calculations | 16 |
| Recruitment | 15 | Strategies for achieving adequate participant enrollment to reach target sample size | NA |

**Methods: Assignment of interventions (for controlled trials)**

Allocation:

| | | | |
|---|---|---|---|
| Sequence generation | 16a | Method of generating the allocation sequence (eg, computer-generated random numbers), and list of any factors for stratification. To reduce predictability of a random sequence, details of any planned restriction (eg, blocking) should be provided in a separate document that is unavailable to those who enroll participants or assign interventions | 10 |
| Allocation concealment mechanism | 16b | Mechanism of implementing the allocation sequence (eg, central telephone; sequentially numbered, opaque, sealed envelopes), describing any steps to conceal the sequence until interventions are assigned | 10 |
| Implementation | 16c | Who will generate the allocation sequence, who will enroll participants, and who will assign participants to interventions | 9-10 |

| | | | |
|---|---|---|---|
| Blinding (masking) | 17a | Who will be blinded after assignment to interventions (eg, trial participants, care providers, outcome assessors, data analysts), and how | __NA__ |
| | 17b | If blinded, circumstances under which unblinding is permissible, and procedure for revealing a participant's allocated intervention during the trial | __N/A__ |

**Methods: Data collection, management, and analysis**

| | | | |
|---|---|---|---|
| Data collection methods | 18a | Plans for assessment and collection of outcome, baseline, and other trial data, including any related processes to promote data quality (eg, duplicate measurements, training of assessors) and a description of study instruments (eg, questionnaires, laboratory tests) along with their reliability and validity, if known. Reference to where data collection forms can be found, if not in the protocol | _10-11 |
| | 18b | Plans to promote participant retention and complete follow-up, including list of any outcome data to be collected for participants who discontinue or deviate from intervention protocols | __10-11, 16-17 |
| Data manageme nt | 19 | Plans for data entry, coding, security, and storage, including any related processes to promote data quality (eg, double data entry; range checks for data values). Reference to where details of data management procedures can be found, if not in the protocol | Supplement 4 |
| Statistical methods | 20a | Statistical methods for analyzing primary and secondary outcomes. Reference to where other details of the statistical analysis plan can be found, if not in the protocol | __16-19__ |
| | 20b | Methods for any additional analyses (eg, subgroup and adjusted analyses) | __17-18__ |
| | 20c | Definition of analysis population relating to protocol non-adherence (eg, as randomized analysis), and any statistical methods to handle missing data (eg, multiple imputation) | _16-17, 19 |

**Methods: Monitoring**

| | | | |
|---|---|---|---|
| Data monitoring | 21a | Composition of data monitoring committee (DMC); summary of its role and reporting structure; statement of whether it is independent from the sponsor and competing interests; and reference to where further details about its charter can be found, if not in the protocol. Alternatively, an explanation of why a DMC is not needed | 15, Supplement 6 |
| | 21b | Description of any interim analyses and stopping guidelines, including who will have access to these interim results and make the final decision to terminate the trial | 15, Supplement 6 |
| Harms | 22 | Plans for collecting, assessing, reporting, and managing solicited and spontaneously reported adverse events and other unintended effects of trial interventions or trial conduct | 10 |
| Auditing | 23 | Frequency and procedures for auditing trial conduct, if any, and whether the process will be independent from investigators and the sponsor | Supplement 3 |

**Ethics and dissemination**

| | | | |
|---|---|---|---|
| Research ethics approval | 24 | Plans for seeking research ethics committee/institutional review board (REC/IRB) approval | 3, 20 |
| Protocol amendments | 25 | Plans for communicating important protocol modifications (eg, changes to eligibility criteria, outcomes, analyses) to relevant parties (eg, investigators, REC/IRBs, trial participants, trial registries, journals, regulators) | Supplement 7 |
| Consent or assent | 26a | Who will obtain informed consent or assent from potential trial participants or authorized surrogates, and how (see Item 32) | 20 |
| | 26b | Additional consent provisions for collection and use of participant data and biological specimens in ancillary studies, if applicable | N/A |
| Confidentiality | 27 | How personal information about potential and enrolled participants will be collected, shared, and maintained in order to protect confidentiality before, during, and after the trial | Supplement 4 |
| Declaration of interests | 28 | Financial and other competing interests for principal investigators for the overall trial and each study site | 22 |

| | | | |
|---|---|---|---|
| Access to data | 29 | Statement of who will have access to the final trial dataset, and disclosure of contractual agreements that limit such access for investigators | Supplement 4 |
| Ancillary and post-trial care | 30 | Provisions, if any, for ancillary and post-trial care, and for compensation to those who suffer harm from trial participation | __N/A |
| Dissemination policy | 31a | Plans for investigators and sponsor to communicate trial results to participants, healthcare professionals, the public, and other relevant groups (eg, via publication, reporting in results databases, or other data sharing arrangements), including any publication restrictions | 21, Supplement 8 |
| | 31b | Authorship eligibility guidelines and any intended use of professional writers | __22 |
| | 31c | Plans, if any, for granting public access to the full protocol, participant-level dataset, and statistical code | Supplement 8 |

**Appendices**

| | | | |
|---|---|---|---|
| Informed consent materials | 32 | Model consent form and other related documentation given to participants and authorized surrogates | __N/A__ |
| Biological specimens | 33 | Plans for collection, laboratory evaluation, and storage of biological specimens for genetic or molecular analysis in the current trial and for future use in ancillary studies, if applicable | __N/A__ |

*It is strongly recommended that this checklist be read in conjunction with the SPIRIT 2013 Explanation & Elaboration for important clarification on the items. Amendments to the protocol should be tracked and dated. The SPIRIT checklist is copyrighted by the SPIRIT Group under the Creative Commons "Attribution-NonCommercial-NoDerivs 3.0 Unported" license.

**2. List of Vanderbilt Learning Healthcare System Platform Investigators**

Vanderbilt University Medical Center and Vanderbilt University, Nashville, TN –

Gordon Bernard, Bob Dittus, Shon Dwyer, Robert Freundlich, Cheryl Gatto, Frank Harrell, Paul Harris, Tina Hartert, Jim Hayman, Catherine Ivory, Ruth Kleinpell, Sunil Kripalani, Christopher Lindsell, Lee Ann Liska, Patrick Luther, Jay Morrison, Thomas Nantais, Jill Pulley, Kris Rehm, Todd Rice[+], Russell Rothman, Matt Semler[+], Robin Steaban, Philip Walker, Consuelo Wilkins, Adam Wright[+], and Autumn Zuckerman.

[+] Denotes members of the ACORN Trial Study Team.

#### 3. Data Auditing

Data manually collected on the front end will be selected from pre-specified selections preventing illogical entries. On the backend, data will go through routine logic and range checking to ensure data quality.

**4. Patient Privacy and Data Storage**

At no time during the course of this study, its analysis, or its publication, will patient identities be revealed in any manner. The minimum necessary data containing patient or provider identities or other private healthcare information (PHI), is collected. All subjects are assigned a unique study ID number for tracking. Data collected from the medical record is entered into the secure online database REDCap. The PHI required to accurately collect clinical and outcomes data is available only to investigators at the site at which the subject is enrolled, and this data is shared only in completely de-identified form with the coordinating center via the secure online database REDCap. Hard copies of the data collection sheet completed at the time of the airway management event are stored in a locked room until after the completion of enrollment and data cleaning. Once data are verified and the database is locked, all hard copies of data collection forms will be destroyed. The de-identified dataset housed in REDCap will be accessed by the coordinating center for reporting the results of this trial. All data will be maintained in the secure online database REDCap until the time of study publication. At the time of publication, all PHI at local centers will be expunged and only the de-identified version of the database will be retained. Potential future use of de-identified data generated in the course of this study by the coordinating center and other participating sites is allowed and will be governed by mutual data sharing use agreements.

#### 5. Definition of Support Therapy Free Days

Days alive and free of the supportive therapy (e.g., delirium and coma-free days, renal replacement therapy-free days, ICU-free days, hospital-free days, ventilator-free days, and vasopressor-free days) will be defined as the number of calendar days alive and free of the supportive therapy from the final receipt of the supportive therapy through 28 days after enrollment [67,68]. The day of enrollment will be considered to be day 0. Outcome ascertainment will cease at the time of hospital discharge or 28 days after enrollment, whichever occurs first. Receipt of the supportive therapy will be considered to end at the time of the patient's final receipt of the supportive therapy between enrollment and 28 days after enrollment. Patients who continue to receive the supportive therapy at day 28 will receive a value of zero. Patients who die prior to day 28 will receive a value of zero. Patients who are discharged from the hospital prior to day 28 and are receiving the supportive therapy at the time of discharge will receive a value of zero. Patients who are removed from the supportive therapy and are discharged from the hospital without the supportive therapy prior to 28 days will be assumed to remain free of the supportive therapy between hospital discharge and day 28. For patients who are removed from the supportive therapy, return to receiving the supportive therapy, and are subsequently removed again from the supportive therapy prior to day 28, days alive and free of the supportive therapy will be counted from the final receipt of the supportive therapy prior to day 28.

#### 6. Data and Safety Monitoring Board Charter

### DATA AND SAFETY MONITORING BOARD CHARTER

***Effect of Antibiotic Choice on ReNal Outcomes (ACORN)***

**December 1, 2021**

**Version 1.0**

**VUMC IRB #210591**

**Edward Qian, MD**

#### TABLE OF CONTENTS


I. DEFINITIONS AND ABBREVIATIONS

II. INTRODUCTION
  i. Scope

III. STUDY OVERVIEW AND STUDY DESIGN

IV. ORGANIZATION OF DSMB MEMBERSHIP
  i. Composition of the DSMB
  ii. Selection of DSMB Members
  iii. DSMB Membership

V. RESPONSIBILITIES AND FUNCTIONS
  i. Overview
  ii. DSMB Responsibilities
  iii. Investigator and Key Personnel Responsibilities
  iv. Unblinded Study Statistician Responsibilities

VI. MEETINGS
  i. DSMB Organizational Meeting
  ii. DSMB Data Review Meetings
  iii. Voting and Meeting Motions
  iv. Meeting Quorum

VII. DSMB REPORT PREPARATION
  i. Data Management
  ii. Generating the DSMB Report
  iii. Defining and Reporting of SAEs

VIII. AMENDMENTS TO THE DSMB CHARTER

IX. CONFIDENTIALITY

X. CONFLICT OF INTEREST

XI. DSMB CENTRAL FILES
  i. DSMB Member File
  ii. DSMB Report File

XII. DSMB MEMBER AGREEMENT

XIII. APPENDIX/ATTACHMENTS
  i. Conflict of Interest Disclosures
  ii. Data and Safety Monitoring Plan
  iii. DSMB Members, Investigators, Key Study Personnel
  iv. Other DSMB Documents

#### I. DEFINITIONS AND ABBREVIATIONS

Safety monitoring: Any process during a clinical study that involves the review of accumulated data for groups of patients to determine if any of the procedures practiced should be altered or stopped because they present a safety risk. Safety monitoring is carried out to ensure protection of human participants while maintaining the scientific integrity of the trial.

Data and Safety Monitoring Board (DSMB): A committee of experts, independent of the trial investigators and funding agency, that periodically reviews the conduct and results of the trial and that makes recommendations regarding continuation without change, continuation with change, or termination of the trial.

DSMB Biostatistician: A voting member of the DSMB, who will provide statistical expertise at DSMB meetings and is responsible for interpretation of data analysis provided to the DSMB.

DSMB Chairperson: A voting member of the DSMB who will chair all DSMB meetings.

Principal Investigator: The investigator who takes overall responsibility for, and is accountable for, all activities of the study.

Study Biostatistician: The biostatistician who is responsible for the Data Management Plan and Statistical Analysis Plan for the study.

Adverse Event (AE)
Conflict of Interest (COI)
Data and Safety Monitoring Board (DSMB)
Data and Safety Monitoring Plan (DSMP)
Good Clinical Practice (GCP)
Institutional Review Board (IRB)

Principal Investigator (PI)
Serious Adverse Event (SAE)
Statistical Analysis Plan (SAP)
Vanderbilt Human Research Protections Program (VHRPP)

#### II. INTRODUCTION

The DSMB provides independent safety review and trial guidance during the course of an ongoing study. The DSMB periodically reviews accruing data and safety evaluations and judges whether the overall safety and feasibility of the trial remains acceptable. This DSMB is responsible for assessing the validity and ongoing safety for this study comparing the effect of anti-pseudomonal cephalosporins versus anti-pseudomonal penicillins on the incidence of acute kidney injury among acutely ill patients. The purpose of this charter is to define the procedures for the DSMB, specify roles and responsibilities, delineate qualifications of the members, describe the purpose and frequency of meetings, provide the procedures for ensuring confidentiality and proper communication, outline the content of the DSMB reports, and detail the formal operating procedures. This DSMB will function in accordance with VHRPP policy and procedures VI.E and VI.E.1.

This charter is a living document, which may be revised before or throughout the conduct of the study by the DSMB and PI. Revision by the DSMB will be subject to the approval of the PI and revision by the PI will be subject to the approval of the DSMB.

**Scope**

This DSMB charter will apply to the protocol for *Effect of Antibiotic Choice on ReNal Outcomes (ACORN)*. For the purposes of this study, any recommendations to alter study conduct will be based on safety and efficacy. A formal interim analysis will occur once 50% patient accrual has been met. Based on the results during the interim analysis, the statistical operating characteristics of the final analysis may be affected. Safety will be assessed by unblinded review of SAEs, unanticipated events, and outcomes. The DSMB will specifically review summary reports of safety data and may review individual cases if deemed appropriate or necessary to determine if a safety concern is emerging. These recommendations will be directed to the PI who has the responsibility to accept, reject or modify DSMB recommendations.

#### III. STUDY OVERVIEW AND STUDY DESIGN

This study aims to examine the impact of anti-pseudomonal cephalosporins versus anti-pseudomonal penicillins in the treatment of sepsis in acutely ill adults. This study has been determined by the IRB to be minimal risk and a waiver of informed consent was approved. The study will include patients 18 years old or greater presenting less than 12 hours to the emergency department and medical intensive care units at Vanderbilt University Medical Center in Nashville, TN for the treatment of sepsis. The PI for the trial, Dr. Edward Qian, is located at Vanderbilt University Medical Center in Nashville, TN**.** See the attached study protocol for complete study details.

#### IV. ORGANIZATION OF DSMB MEMBERSHIP

**Composition of the DSMB**

The DSMB will consist of three voting members, independent of the study team, with collective expertise relevant to this study and study population. Membership of the DSMB will reflect the disciplines and medical specialties necessary to interpret the data from this trial, including biostatistics, infectious disease, critical care, and clinical trials. The DSMB will include an independent biostatistician. One DSMB member will be appointed the DSMB chairperson.

**Selection of DSMB Members**

The three DSMB members will be initially selected by the PI in conjunction with the Learning Healthcare System (LHS) Platform. The voting members will elect the DSMB chairperson from among the members. In the event a DSMB member is unable to complete their duties, the PI and remaining DSMB members will identify a suitable replacement. If the member unable to complete their duties is serving as the DSMB chairperson, a new chairperson will be elected by all three members of the DSMB after the replacement has been incorporated into the DSMB.

**DSMB Membership**

DSMB membership will be voluntary. Members must report potential COI and be cleared of any real or potential conflicts of interest in accordance with provisions in this charter.

#### V. RESPONSIBILITIES AND FUNCTIONS

**Overview**

The ongoing review of data by an independent committee provides reassurance to the investigator(s) that the clinical study does not jeopardize participant safety while ensuring the scientific integrity of the trial. The DSMB will be independent of the study team and will conduct a formal interim analysis once 50% patient accrual has been met, and *ad hoc* as needed. During these confidential meetings, the DSMB will assess subject safety and make recommendations.

**DSMB Responsibilities**

The DSMB shares responsibility with the PI for monitoring the safety of participants, and for ensuring that participants are not exposed to undue risk. Following DSMB meetings, the DSMB will provide recommendations regarding the continuing safety, validity, and scientific integrity of the study to the PI. The DSMB chairperson may verbally communicate recommendations and/or areas of concern related to the study at the end of each DSMB meeting to the PI, or designee. A written copy of the recommendations will be drafted, and the recommendations will be signed by the DSMB chairperson and provided to the PI within 10 business days of the DSMB meeting. The following recommendations can be made:

- Continuation of the trial without modification.
- Continuation of the trial with specified modifications (modifications may include, but are not limited to, changes in inclusion/exclusion criteria, and alterations in study procedures.)
- Termination of the trial.

Acting independently, without interest in influencing the outcome of the study, DSMB members are responsible for:

- Reviewing and approving this DSMB charter and plans for data and safety monitoring prior to study start.
- Attending pre-specified and *ad hoc* DSMB meetings.
- Evaluating the progress of the trial, including periodic assessments of data quality and timeliness, accrual, participant risk versus benefit, and other factors that can affect study integrity.
- Identifying, with input from the PI, potential unintended consequences of actions and decisions made by the DSMB or the study team.
- Considering factors external to the study when relevant information becomes available, such as scientific or therapeutic developments that may have an impact on the safety or the ethics of the trial.
- Making recommendations to assist in the resolution of problems reported by the PI, relevant parties or identified by the DSMB.
- Ensuring the confidentiality and blinding of study data when appropriate.
- Recording and communicating meeting minutes and appropriate reports to necessary parties.

**Investigator and Key Personnel Responsibilities**

The PI or designee is responsible for the coordination of the DSMB communications, activities, and materials including the following:

- Recommending DSMB members and providing the initial draft of the DSMB Charter.
- Managing any transfer of the clinical safety data.
- Preparing and validating periodic reports containing summaries of the safety data pertinent to DSMB review as outlined in the attached DSMB report templates.
- Distributing open reports to the DSMB prior to the DSMB meeting.
- Coordinating distribution and collection of the closed DSMB report with the study statistician or a designee.
- Documenting and communicating meeting minutes in appropriate reports to necessary parties.
- Preparing and distributing *ad hoc* reports.
- Scheduling DSMB meetings and conference calls and preparing and distributing agendas under the direction of the DSMB Chairperson.
- Preparing summary minutes for the open portion of each DSMB meeting and maintaining all open meeting records.
- Maintaining the DSMB files and archives of electronic datasets and programs used to generate each summary report.
- Making resources available in a timely fashion to the DSMB as required to carry out its designated functions including:
    - Study documents (protocols and amendments)
    - Study data
    - SAE reports
    - Additional medical records and supporting documentation as requested to address specific safety concerns
    - Other data as requested in writing by the DSMB

**Unblinded Study Statistician Responsibilities**
An unblinded statistician or designee is responsible for coordinating closed meetings of the DSMB, including communications, activities, and materials.

- Preparation of open and closed DSMB reports.
- Distribution of the closed DSMB report (and collection of the reports if distributed in hard copy).
- Receiving and maintaining copies of closed session minutes.
- Maintaining the blind for all study team members.

#### VI. MEETINGS

Scheduled DSMB meeting types include organizational and data review meetings. DSMB meetings may be conducted face-to-face or virtually. All DSMB meeting discussions are considered confidential. The DSMB will meet once at study start and then during a formal interim analysis, which will occur once 50% patient accrual is met. The PI or designee will send DSMB members a study report and meeting agenda at least one week prior to each scheduled meeting. The unblinded study statistician or designee will provide a closed report for review by the DSMB at least one week prior to each scheduled meeting. Unscheduled meetings include *ad hoc* meetings requested in writing by the DSMB, the PI, or other authorized person to look at specific data if considered necessary to ensure the integrity of the study and safety of participants.

**DSMB Organizational Meeting**
The DSMB will meet at study start for an organizational meeting. The organizational meeting agenda will resemble the following:

- Operational Plan (DSMB charter)
    - Roles and responsibilities
    - The structure of meetings (open and closed sessions)
    - Discussion and voting procedures
    - Reporting to and from the DSMB
- Protocol (IRB approved)
    - Background/rationale
    - Primary and secondary objectives
    - Outcome measures
    - Study design
    - Study procedures
    - Study timeline
- Data and safety monitoring plan
    - Adverse event system and unintended outcomes
    - Content of interim reports (safety outcomes of interest)
    - Procedures to manage and access reports
    - Frequency of interim reports

**DSMB Data Review Meetings**
The PI and other investigators will monitor the data on a daily basis for accuracy, completeness, and adverse events. The DSMB will be responsible for reviewing adverse events and conducting an interim analysis. They will be asked to be available for rapid access by the investigators in the case of the need to evaluate serious and unexpected adverse events or any other major unanticipated or safety related issues. Furthermore, in cases of serious and unexpected adverse events, the DSMB will have the ability to pause the trial to investigate possible safety issues and/or suggest changes to the design of the study to abrogate any safety issues.

The DSMB will conduct a single interim analysis for efficacy and safety at the anticipated halfway point of the trial; once 50% patient accrual (1025 patients) has been met. The stopping boundary for efficacy will be met if the P value for the difference between groups in the primary outcome is 0.001 or less. Given the minimal risk nature of the study and current use of both interventions as a part of usual care, there will be no stopping boundary for futility. At the interim analysis, the DSMB will also monitor the distribution of the acute kidney injury (AKI) ordinal outcome within the anti-pseudomonal penicillin group and may propose to increase the planned sample size to maintain the pre-planned power to detect an odds ratio of 0.65. Additionally, the DSMB will reserve the right to stop the trial at any point, request additional data or interim analyses, or request modifications of the study protocol as required to protect patient safety. The DSMB may modify the frequency of these data review meetings at any time. The DSMB chairperson, in consultation with the other voting members and the PI, will be responsible for revising the type and frequency of DSMB data review meetings.

The data review meeting agenda will resemble the following:

- Open session - trial performance
    - Accrual

  - Retention
  - Protocol violations
  - Summary of any monitoring reports
  - Baseline characteristics
- Move to closed session
  - Review unblinded summary safety data
- Voting DSMB members makes recommendations
  - Continuation of trial without modification
  - Continuation of trial with specified modifications

**Voting and Meeting Motions**

All motions must be approved by a majority vote of the DSMB voting members. Every DSMB voting member will have one vote per recommendation to be submitted to the PI. To vote, a DSMB member must be present at convened scheduled meetings or participate through teleconference, webinar, e-mail, etc. A simple majority of members present passes a proposal, motion, or recommendation to the PI. Only the recommendation of study termination requires a unanimous DSMB vote. The DSMB chairperson should provide such a recommendation to the PI or designee immediately.

**Meeting Quorum**

This DSMB is comprised of three experts; all three members must be present to establish a quorum. A quorum is required for any meeting.

#### VII. DSMB REPORT PREPARATION

**Data Management**

For each DSMB meeting involving data review, a cut-off date for receipt of data to be included in the DSMB report will be established. The data cut-off date will be specified in the report prepared for the DSMB.

**Generating the DSMB Report**

The study biostatistician or designee will develop a draft DSMB Report. The report will be based upon the safety responsibilities of the DSMB. The report will contain a brief discussion of the methods and analyses as well as table shells, listing shells, and graphical mockups. The report will be reviewed and agreed upon by the PI, the study biostatistician, and the DSMB. The DSMB members may at any time request additional data, report format modifications or changes to the reporting timelines when necessary, including for emergency meetings. Interim statistical analyses to compare arms will not be conducted, so monitoring of the study will not affect the statistical operating characteristics of the final analysis.

**Defining and Reporting of SAEs**

Subjects in this study are acutely ill patients receiving anti-pseudomonal cephalosporins or anti-pseudomonal penicillins in the treatment of sepsis. As such, subjects may experience adverse events related to the administration of anti-pseudomonal cephalosporins or anti-pseudomonal penicillins. These events are anticipated and will be captured as part of routine clinical care and are not considered safety events. All study interventions are within the spectrum of standard of care. Patients are evaluated at least daily at the bedside and screened for side effects of anti-pseudomonal cephalosporins or anti-pseudomonal penicillins on daily rounds. Patients will be excluded from the study if a current documented allergy to anti-pseudomonal cephalosporins or anti-pseudomonal penicillins is present or if the treating clinician feels that either an anti-pseudomonal cephalosporin or anti-pseudomonal penicillin is required or contraindicated for the optimal treatment of the patient, including a more directed antibiotic therapy against known prior resistant infections or suspected sepsis with an associated central nervous system infection. All decisions regarding management of patient care will be at the discretion of the treating provider and will not be restricted or altered in any way by the study. The Vanderbilt IRB has determined that this trial poses no greater than minimal risk to study subjects, and SAEs or other problems are not anticipated.

Safety reporting will include those events identified *a priori* which may be relevant to the DSMB's consideration of the risk/benefit profile of the study. These are unrelated to the research itself but are known to be related to the therapies under evaluation, such as anaphylaxis. This and any other AE that is unexpected and related or possibly related to the research will be reported to the IRB and the DSMB. In the unlikely event that such events occur, serious unanticipated problems involving risks to subjects or others will be reported immediately as appropriate. Adverse events will be reported within 3 days to the PI, the PI will then report to the DSMB & IRB within 7 days, and a written report will be sent to the DSMB and IRB within 15 calendar days. The PI will apprise fellow investigators and study personnel of all such events that occur during the conduct of this research project through regular study meetings. Annual reports will be submitted by the study coordinator to the responsible IRBs summarizing study progress, AEs, complaints about the research, and any protocol violations.

#### VIII. AMENDMENTS TO THE DSMB CHARTER

This DSMB Charter can be amended as needed during the study. All amendments will be documented with a new version number and dates and will be recorded in the minutes of the DSMB meeting. Each revision will be reviewed and agreed upon by the PI and the DSMB.

#### IX. CONFIDENTIALITY

All committee members will treat as confidential the reports, meeting discussions, minutes, and any other data provided for review by the DSMB. Data prepared for the DSMB should not be shared by the DSMB with any person outside of or involved in the conduct of the study. Master copies of the DSMB reports and recommendations will be maintained in a limited access environment (e.g. locked cabinet, or secure server with user level access control).

#### X. CONFLICT OF INTEREST

DSMB members will follow standard COI guidelines and be cleared of any real or potential COIs. DSMB members should remain separate and independent from the study. Members of the DSMB must have no direct interest, financial or otherwise, in knowing or influencing the outcome of the study. DSMB members should be mindful of relationships with trial investigators that could be considered reasonably likely to affect their objectivity. DSMB members should not be in a position of decision making regarding the care of patients who are participating in the study. Prior to confirmation as a DSMB member, each potential voting member will disclose any potential COI and complete a COI statement. Once this form has been signed by the potential DSMB member, it will be forwarded to the PI for review and signature. When the signatures of the DSMB members and the PI have been obtained, one copy of the form will be maintained by the PI or designee for safekeeping. DSMB members are responsible for advising the PI of changes in their COI Statement. Members will be asked to confirm that they have no new COI prior to each DSMB meeting. Members who develop significant or potentially significant COI will be required to resign from the DSMB.

#### XI. DSMB CENTRAL FILES

The documents listed below will be maintained by the PI or designee and made available to the DSMB and appropriate IRB upon request.

**DSMB Member File**

The PI or designee is responsible for maintaining a folder that contains for each voting DSMB member:

- DSMB member's name and contact information
- DSMB member's CV
- Signed Confidentiality Agreement
- Signed Conflict of Interest Statements (initial and annual)
- Signed versions of the DSMB Charter
- Copies of communication between the study team and DSMB members

**DSMB Report File**

The study biostatistician or a designee will be responsible for maintaining a file of written documents, which will include but may not be limited to the following:

- Closed session reports
- Closed session meeting minutes

#### XII. DSMB MEMBER AGREEMENT

As a voluntary member of the Data and Safety Monitoring Board (DSMB) for the evaluation of the effects of anti-pseudomonal cephalosporins versus anti-pseudomonal penicillins on the incidence of acute kidney injury among acutely ill patients, I agree to adhere to the terms and procedures described in this charter. I confirm that I have read this DSMB charter; I understand it, and I will work according to this charter and to the ethical principles stated in the latest version of the applicable guidelines for GCP, or the applicable laws and regulations, as appropriate.

____________________________________________ ____________________

*Full Name, Title, Role/Department* *Date*

**7. Plan for communication of protocol changes**

Any changes to the trial protocol (e.g., changes to eligibility criteria, outcomes, analyses) will be implemented via a new version of the full trial protocol, tracked with the date of the update and the version number of the trial protocol. A list summarizing the changes made with each protocol revision will be included at the end of each protocol. The updated protocol will be sent to the relevant IRBs for tracking and approval prior to implementation of the protocol change. At the time of publication, the original trial protocol, and the final trial protocol, including the summary of changes made with each protocol change, will be included in the supplementary material for publication.

#### 8. Public Access

Data will be made available to researchers whose research proposal is approved by the principal investigator in addition to approval by an Institutional Review Board and an executed data use agreement. Data will become available 3 months following publication of outcomes and will remain available for at least 5 years.
